# Effectiveness of educational intervention on breast cancer knowledge and breast self-examination among female university students in Bangladesh: a pre-post quasi-experimental one group study

**DOI:** 10.1101/2021.10.20.21265265

**Authors:** Rumpa Sarker, Md. Saiful Islam, Mst. Sabrina Moonajilin, Mahmudur Rahman, Hailay Abrha Gesesew, Paul R Ward

## Abstract

**Background:** Breast cancer is a global health issue and a leading cause of death among women. Early detection through increased awareness and knowledge on breast cancer and breast cancer screening is thus crucial. The aim of the present study was to assess the effect of educational intervention program on breast cancer knowledge and practice of breast self-examination among young female students of a university in Bangladesh.

**Methods:** A quasi-experimental one group (pre-post) study design was conducted at Jahangirnagar University in Bangladesh. Educational information on breast cancer and breast self-examination (BSE), demonstration of BSE procedure and leaflets were distributed among 400 female students in common room setting in dormitories after obtaining written informed consent. The stepwise procedures of BSE performance were demonstrated with images. Pre-intervention and 15 days post-intervention assessments were conducted to assess the changes in knowledge on breast cancer and practices of BSE. Mc-Nemar’s tests and paired sampled *t*-tests were performed to investigate the differences between pre- and post-test stages.

**Results:** Significant changes were found in knowledge and awareness about breast cancer and BSE practices after the educational session. The significant differences were measures in the mean scores of pre-test vs. post-test: breast cancer symptoms (2.99±1.05 vs. 6.35±1.15; *p*<0.001), risk factors (3.35±1.19 vs. 7.56±1.04; *p*<0.001), treatment (1.79±0.90 vs. 4.63±0.84; *p*<0.001), prevention (3.82±1.32 vs. 7.14±1.03; *p*<0.001), screening of breast cancer (1.82±0.55 vs. 3.98±0.71; *p*<0.001) and process of breast self-examination (1.57±1.86 vs. 3.94±0.93; *p*<0.001). Likewise, a significate percentage of change in BSE practices was obtained between pre-test and post-test (21.3% vs. 33.8%; *p*<0.001)

**Conclusions:** This study findings confirm that the study population had poor awareness and knowledge at baseline that was improved significantly after educational session. A nationwide reach-out with community-based interventions is recommended for female population in both rural and urban areas.

## Introduction

Breast cancer is considered as worldwide health concern and one of the prominent causes of mortality among women. In 2018, approximately 2 million new cancer cases were detected that is projected as 23% of all cancers, the most occurring cancer among women [1]. In Bangladesh, breast cancer is ranked as the second most leading cancer after cervical carcinoma and in females, these two cancers constitute 38% of all cancers [2]. This continuously rising burden has been a matter of concern for a long time now, especially for limited resourced countries like Bangladesh [3]. As curative treatment for any cancer is yet not available, several approaches have been advocated towards increasing awareness that may lead to early detection of cancers including breast cancer [4]. Breast cancer education and awareness in limited resourced countries can be a key to initiate the early detection of breast cancer and subsequently increase the survival rate [5]. Breast abnormalities can be detected by own in many cases as it occurs in a visible organ, and enables to seek medical assistance as soon as possible. For this, women must know how their own breast should look and feel normally and also be aware of all the danger signs, as suggested by the American Cancer Society [6].

Nemours findings suggest that educating the community about assessment of asymptomatic women has the potential to increase the proportion of breast cancer detected at an early stage. Studies conducted on female students of Turkey, Malaysia, Ahmedabad have showed significantly improved knowledge and awareness on breast cancer after educational session using various health educational tools such as group discussion sessions, video demonstration, pamphlet, etc. [7-9]. Findings from a pilot mobile intervention program in Bangladesh has reported that in comparison with control group, the women who attended to an educational intervention were more likely to visit to clinics for a follow-up to check for abnormalities found in their breast examination [10]. This strengthens the vital role of education in decreasing late presentation.

Recommended screening methods like Mammogram, clinical visit for clinical breast examination, ultrasound and MRI are expensive for a large population group in low-income socioeconomic setup and whole nationwide screening program is not feasible. Moreover, lack of knowledge and awareness about breast cancer has been reported from some studies conducted on females of Bangladesh that may contribute to less adherences of women to receive recommended screening [11-14]. Therefore, creating breast cancer awareness through educational intervention among females can be a feasible solution to early detection. This is only possible, if we know the present level of knowledge, attitude and practices of the female population towards breast cancer. The currently available data is limited to some sections of the society and related to few aspects of the disease. This study was planned to assess the knowledge and practice level of breast cancer and breast self-examination (BSE) among female respondents (pre-test) and to note the changes after educational session in knowledge of women about ‘risk factors, symptoms, diagnosis, and treatment modalities of breast cancer, and to know about practice of BSE (pre-test vs. post-test) in females and to find out the effectiveness of the educational session on the respondents. The young female university students aged from 18-26 are already passing their reproductive age and are the future mothers. Also, the study population of our study is considered to be the most educated segment of the population. The assessment of their current knowledge and practice of breast cancer is crucial. Besides, to educate women about the warning sign symptoms and strive for improvement of health seeking behavior by making them aware is an important step to drag down high incidence and mortality rate from breast cancer. Additionally, creating awareness, knowledge about breast cancer and increased practice of breast cancer among them may have double benefits. Firstly, it may create positive impact and increase the awareness about breast cancer. Secondly, as they belong to the most educated population, they can help in spreading the knowledge and awareness among their own family, friends and community in large.

## Methodology

### Study design and setting

A one-group pre-post quasi-experimental interventional study was conducted among female university students residing in dormitories of Jahanginagar University in Dhaka, Bangladesh from December 2019 to March 2020. Jahanginagar University is the largest and only one fully residential university in Bangladesh.

### Participants and procedures

The study was conducted in 400 female respondents of aged 18-26 years corresponding to Honors 1^st^ year to master’s students. Inclusion criteria included: being female students residing in the university’s dormitories and being aged from 18-26 years old. Exclusion criterion was being female students who didn’t reside within residential dormitories. The proportionate stratified random sampling technique was conducted to calculate the study sample from each dormitory. In this approach, each stratum sample size was directly proportional to the population size of the entire population of strata. The study was carried out in three phases: first phase (pre intervention phase), second phase (intervention phase) and third phase (post intervention phase).

#### First phase (pre-test phase)

A pre-designed structured interview questionnaire was used to collect the following data from the respondents: socio-demographic data, respondent’s knowledge, attitude and practice regarding breast cancer, screening, and BSE.

#### Second phase (the intervention phase)

Materials (e.g., lecture/discussions, brain storming, leaflet where the stepwise process of BSE was documented with images, etc.) were used during the interventional phase. All the sessions were conducted in the respective dormitories of the respondents. Participants were divided into very small groups approximately 10-15 people per group to conduct the sessions so that the educational intervention could be clearly understood by the participants. Each session took 45-60 minutes. After the pre-test session, the participants were given a short break to rearrange themselves into divided groups and get prepared for the educational session. Both pre-test and intervention sessions were conducted on the same day. Each participant was assigned a unique ID number so that they could be traced back for the post-test session. To ensure that the respondents could understand the educational materials, in every session one or two respondents from each group were encouraged to demonstrate and share what they had learned. This was also chosen randomly among the participants who were willing to perform.

#### Third phase (post-test phase)

After 15 days of the conduction of the health education program, participants were traced back according to their IDs. The participants could be traced back successfully and 100% response rate was obtained. This was understandable because all the respondents of this study were the residential students of the university who were residing at their own dormitories during the full period of study. Also, the participants were enough liable to their commitment to fulfill the study while giving consent in the first place. Still, some participants were not present for the post-test session. To trace them back we had to contact them via their contact numbers personally which was obtained from them with their full informed consent by assuring the confidentiality and thus got the post-test data from all of the participants. During third-phase or post-test phase, they were exposed to the same preliminary questions in the pre-test questionnaire to assess the changes in knowledge level and evaluation of the effectiveness of the developed educational program was made.

### Study instruments

A pre-tested and semi-structured questionnaire including informed consent, socio-demographic information and questions related to knowledge towards breast cancer, and BSE practices, was prepared for the study through extensive literature review [15-18].

The questionnaire was reviewed by an external reviewer who was an oncologist and had sound knowledge about breast cancer. Likewise, a pilot test was conducted to assess the readability of the questionnaire. The questionnaire was finalized after incorporating minor amendments based on the participants’ feedback during the piloting periods. A paper-pen-based survey was conducted among participants. Additionally, few post-test data were obtained via telephone from the respondents who couldn’t be present during post-test session for their own personal reasons.

#### Socio-demographic information

Socio-demographic information was recorded during the survey including age, study year (1^st^/2^nd^/3^rd^/4^th^/Master’s), marital status (unmarried/unmarried), family history of breast cancer (yes/no), and relationship with breast cancer affected patient (mother/sister/cousin/aunt/grandmother).

#### Knowledge towards breast cancer measures

To assess the participants’ knowledge towards breast cancer, a total of 43 questions regarding breast cancer (i.e., 8 for symptoms, 10 for risk factors, 6 for treatment, 8 for prevention, 5 for screening, and 5 for process of BSE) were asked during the survey. All questions were answered with three possible responses (i.e., yes/no/don’t know). The distributions of all questions (both pre-test and post-test) are presented in **Additional file**.

#### BSE practices

A single construct (i.e., *Have you ever self-examined your breast for breast cancer?*) was used to assess the BSE with binary responses (yes/no).

### Statistical analysis

The SPSS version 25.0 was used for processing and analyzing data. Descriptive statistics were performed. To assess the differences between pre-test and post-test, Mc-Nemar tests and paired sample t-tests were computed as appropriate. Before performing the Mc-Nemar tests, each question was transformed into dichotomous (i.e., correct answer and wrong answer). A *p*-value less than 0.05 was deemed as statistically significant.

### Ethical consideration

The study was conducted in accordance with the Institutional Research Ethics guidelines and ethical principle involving human participation (i.e., Helsinki Declaration). Formal ethics approval was granted by the Biosafety, Biosecurity, and Ethical Clearance Committee, Jahangirnagar University, Savar, Dhaka-1342, Bangladesh. At first, all participants were informed about the purpose and objectives of the study. Participants were informed that it was a three-phase study, and also about the duration of the study and the approximate time that would be taken from them. Then, written informed consents were taken from each of them who agreed to participate in the study. All information related to participants were kept confidential.

## Results

### General characteristics of participants

The sample comprised of a total of 400 female university students aged between 18-26 years. Of them, majority were 18-21 years old (35.3%), students of 2^nd^ year (23.5%), and most were unmarried (86%) (Table 1). In terms of a family history of breast cancer, 18.3% participants reported that someone in their family had been diagnosed with the disease which included mother (11.6%), sister/cousin (24.6%), aunt (40.6%), and grandmother (23.2%). The remaining 81.2% had no family history of breast cancer.

**Table 1:** Socio-demographic variables

| **Variables** | **n (%)** |
| --- | --- |
| **Age** |  |
| 18-20 | 141 (35.3) |
| 21-23 | 124 (31.0) |
| 24-26 | 135 (33.8) |
| **Study year** |  |
| 1st | 78 (19.5) |
| 2nd | 94 (23.5) |
| 3rd | 87 (21.8) |
| 4th | 84 (21.0) |
| Master's | 57 (14.2) |
| **Marital status** |  |
| Unmarried | 344 (86.0) |
| Married | 56 (14.0) |
| **Family history of breast cancer** |  |
| Yes | 73 (18.3) |
| No | 327 (81.8) |
| **Relationship with affected patient** |  |
| Mother | 8 (11.6) |
| Sister/cousin | 17 (24.6) |
| Aunt | 28 (40.6) |
| Grandmother | 16 (23.2) |

### Effectiveness of intervention on breast cancer knowledge and BSE

Table 2 presents the overall differences in the participants’ knowledge regarding symptoms, risk factors, treatment, prevention, screening methods and process of BSE examination. Significant knowledge differences in the mean scores were obtained between pre-test and post-test: breast cancer symptoms (2.99 ± 1.05 vs. 6.35 ± 1.15; *p* < 0.001), risk factors (3.35 ± 1.19 vs. 7.56 ± 1.04; *p* < 0.001), treatment (1.79 ± 0.90 vs. 4.63 ± 0.84; *p* < 0.001), prevention (3.82±1.32 vs. 7.14±1.03; *p* < 0.001), screening of breast cancer (1.82 ± 0.55 vs. 3.98 ± 0.71; *p* < 0.001) and process of breast self-examination (1.57 ± 1.86 vs. 3.94 ± 0.93; *p* < 0.001). Likewise, a significate percentage of change in BSE practices was obtained between pre-test and post-test (21.3% vs. 33.8%; *p* < 0.001) (Table 3). The distribution and changes of the participants’ knowledge regarding symptoms, risk factors, treatment, prevention, screening methods and process of BSE examination are presented in **Additional file** (Table S1-S5).

**Table 2:** Assessment of total difference in the knowledge of breast cancer among participants (pre-test vs. post-test)

| Variables | Mean | SD | *t*<sup>†</sup> | *p*-value |
| --- | --- | --- | --- | --- |
| **Knowledge about symptoms of breast cancer (total score=8)** |  |  |  |  |
| Post-test | 6.35 | 1.15 | 42.343 | <0.001 |
| Pre-test | 2.99 | 1.05 |  |  |
| Differences | 3.36 | 1.59 |  |  |
| **Knowledge about risks of breast cancer (total score=10)** |  |  |  |  |
| Post-test | 7.56 | 1.04 | 52.161 | <0.001 |
| Pre-test | 3.35 | 1.19 |  |  |
| Differences | 4.21 | 1.62 |  |  |
| **Knowledge about treatment of breast cancer (total score=6)** |  |  |  |  |
| Post-test | 4.63 | 0.84 | 45.211 | <0.001 |
| Pre-test | 1.79 | 0.90 |  |  |
| Differences | 2.84 | 1.26 |  |  |
| **Knowledge about prevention of breast cancer (total score=8)** |  |  |  |  |
| Post-test | 7.14 | 1.03 | 37.350 | <0.001 |
| Pre-test | 3.82 | 1.32 |  |  |
| Differences | 3.32 | 1.78 |  |  |
| **Knowledge about screening of breast cancer (total score=5)** |  |  |  |  |
| Post-test | 3.98 | 0.71 | 47.241 | <0.001 |
| Pre-test | 1.82 | 0.55 |  |  |
| Differences | 2.16 | 0.92 |  |  |
| **Knowledge about process of breast self-examination (total score=5)** |  |  |  |  |
| Post-test | 3.94 | 0.93 | 23.653 | <0.001 |
| Pre-test | 1.57 | 1.86 |  |  |
| Differences | 2.37 | 2.00 |  |  |

Note: <sup>†</sup>Paired t-test; SD= Standard deviation

**Table 3:** Assessment of percentage of changes in practice of Breast self-examination (pre-test vs post-test)

| Variables | Pre-test |  | Post-test |  | Percentage of correct changes | Mc-Nemar test*<br>*p*-value |
| --- | --- | --- | --- | --- | --- | --- |
|  | n | % | n | % |  |  |
| Breast self-examination practice |  |  |  |  |  |  |
| Yes | 85 | 21.3 | 135 | 33.8 | 12.5 | <0.001 |
| No | 315 | 78.8 | 265 | 66.3 |  |  |

## Discussion

The present study found a significant change of knowledge to breast cancer and BSE practices following educational intervention among undergraduate female students in Bangladesh. After educational session and 15 days interval, we assessed the knowledge level of the same respondents. Correct answers were delivered by majority of the respondents about each question in the post-test session. This is consistent with several studies conducted in Egypt, Iran, İzmir (a city of Turkey), and Sivas (a city in the Central Anatolia of Turkey) [19-22]. In these studies, the knowledge level on breast cancer symptoms, risk factors, treatment, prevention, screening methods and practice of BSE were significantly increased after educational session among the respondents. Yilmaz et al. have showed that the mean knowledge score for correct risk factors and correct screening methods were increased to 3.65 ± 2.86 to 9.37 ± 3.10 (total score 12) and 5.45 ± 1.98 to 8.10 ± 1.19 (total score 6), respectively from pre-test to post-test and it supports out finding [22]. Aziz et al. and Rezaein et al. have also revealed in their studies about significant increase in correct knowledge changes about symptoms, risk factors, prevention, and early detection of screening methods after a successful educational/training intervention [19, 20]. The changes in the knowledge level were significant in a previous study by Ceber et al. and correct percentage of changes was higher in the post-test [21]. These findings confirm the significant improvement in knowledge about these segments of breast cancer in the post-test from the pre-test.

In the present study, the knowledge of BSE also increased significantly. The mean difference in the knowledge about process of BSE (total score=5) was 2.37±2.00 (*p*<0.001). This finding is consistent with a previous study by Ceber et al where the knowledge level on BSE were higher in the experimental group who received educational session (control 6. 13 ± 0.91 vs. experimental 7.03 ± 0.84) [21]. The study by Aziz et al. in his study on Egyptian women align with our finding where significant improvement in the knowledge on BSE was found higher after educational session where the mean difference from per-test to post-test in the knowledge about process of breast self-examination was 0.71 ± 0.46 [19]. Another study on female students on the effect of BSE education in Turkey by Beydağ et al. has yielded similar result where the knowledge on BSE was significantly increased. The knowledge level score was 43.2 ± 10.6 before and 68.4 ± 10.5 after the BSE education (*p* < 0.05) [23]. These studies support out finding on the increased knowledge level on BSE. With regard to the practice of BSE, it is found that significant changes from 21.3% to 33.8% (*p* < 0.001) after receiving educational session (Table 3). Similarly, plethora of studies has revealed similar result with this study. In a study in Yazd University, Iran, it was found that before training 62.86% of the women did not perform BSE but, after training this decreased to 33.57% (*p* < 0.001) [24]. Similarly, Ozturk et al. have revealed in his study the ratio of subjects who regularly performed BSE in the study group had increased from 19.0% to 61.3% while the same ratio was found 39.7% in the control group [25]. The differences between two groups and in the same group before and after training were statistically significant [25]. Also, 70.0% did not practice BSE in pre-test compared to 75.0% practicing it in post-test after the intervention, mean practice score was significant (*t* = 9.84, *p* < 0.001) was found in a study by Aziz et al. [19]. All these findings align with this study where the changes in the practice of BSE were significantly increased after the educational session.

Given the fact that, this study was conducted on university students whose educational level were from 1^st^ year of under graduation to post graduation in combination with an efficient, flexible and attractive educational session, this is justifiable that they understood the importance of the information provided at educational session on breast cancer and practice of BSE easily. Thus, all the positive changes in the knowledge and practice of breast cancer and BSE was significantly higher from pre-test to post-test indicating the successful outcome of the educational session that was conducted in this study.

### Limitations

The study had only 15 days interval between the pre-test, educational session and the post-test assessment. So, there’s a big chance of recall bias. If more time interval could be given, that might have impacted the outcomes. No follow-up session of the educational intervention on breast cancer and BSE could be given, also due to time constraints. Respondents were given no reminder to practice breast self-examination during the interval phase. All the data were self-reported by the respondents and no verification could be done to assess the accuracy of the data given by respondents whether the claim of practicing BSE were true or not. The quality of BSE practice could not be assessed. So, it is unknown that if the respondents who are claiming to actually practicing BSE were being able to do it properly or not.

## Conclusion and Recommendation

This educational intervention study was conducted on female university students representing the highly educated segment of the population through a pre- and post-test setting. Thus, the study findings indicated an insight of the effectiveness of educational intervention among university female students regarding breast cancer and BSE focused on awareness and knowledge. The findings indicated that the student’s knowledge regarding breast cancer warning symptoms, risk factors, treatment, prevention, effective screening methods and practice of BSE were very inadequate in the baseline. Hence, this educational intervention used discussion, brain storming and leaflet providing clear, precise and required information about breast cancer and steps for performing BSE had been found to be useful. The results of the post-test of this study suggest that women’s knowledge was significantly increased. However, educational sessions should be continued because increased knowledge level is important to change behavior about early diagnosis for breast cancer. This study concludes that the educational program on breast cancer and BSE had been effective in improvement of knowledge and practice levels of women. Hence, approaches to educational intervention should be given importance. Each and every woman regardless of their socio-demographic conditions need to be educated about breast cancer and breast cancer screening methods. This education should be culturally appropriate and targeted towards individual population so that it can create greater impact. A future study with larger and diversified population is recommended to assess the actual effectiveness and monitor the changes in awareness and practice of breast cancer and breast cancer screening. Also, the focus on the development and implementation of such educational measures to improve breast cancer awareness and practice of screenings among all women in Bangladesh are also needed.

## Data Availability

All data produced in the present work are contained in the manuscript.

## List of abbreviations

BSE: Breast self-examination

## Declaration

### Ethics approval and consent to participate

The study was conducted in accordance with the Institutional Research Ethics guidelines and ethical guidelines involving human participation (i.e., Helsinki Declaration). Formal ethics approval was granted by the Biosafety, Biosecurity and Ethical Clearance Committee, Jahangirnagar University, Savar, Dhaka-1342, Bangladesh. Written informed consent was obtained from all participants.

### Consent for publication

Not applicable.

### Availability of data and materials

The datasets used and/or analyzed during the current study are available from the corresponding author on reasonable request.

### Authors’ contribution

Conceptualization: R.S., M.S.I., M.S.M., M.R., Methodology: R.S., M.S.I., M.S.M., M.R., Investigation: R.S., M.S.M., M.R., Data curation: R.S., M.S.I., Analysis and interpretation of data: M.S.I., Drafting of the manuscript: R.S., M.S.I., Editing: M.S.I., M.S.M., M.R., H.A.G., P.R.W., Critical revision of the manuscript: M.S.I., H.A.G., P.R.W., Supervision: M.S.M.

## Acknowledgement

The authors would like to express the most profound gratitude to all of the respondents who participated in this study.

## References

1. Zaidi Z, Dib HA: The worldwide female breast cancer incidence and survival, 2018. In.: AACR; 2019.

2. Hussain SA, Sullivan R: Cancer control in Bangladesh. Japanese journal of clinical oncology 2013, 43(12):1159–1169.

3. Hussain SMA: Comprehensive update on cancer scenario of Bangladesh. South Asian journal of cancer 2013, 2(04):279–284.

4. Smith RA, Caleffi M, Albert US, Chen TH, Duffy SW, Franceschi D, Nyström L, Detection GSE, Panel AtC: Breast cancer in limited-resource countries: early detection and access to care. The breast journal 2006, 12:S16–S26.

5. El Saghir NS, Adebamowo CA, Anderson BO, Carlson RW, Bird PA, Corbex M, Badwe RA, Bushnaq MA, Eniu A, Gralow JR: Breast cancer management in low resource countries (LRCs): consensus statement from the Breast Health Global Initiative. The Breast 2011, 20:S3–S11.

6. Smith RA, Saslow D, Sawyer KA, Burke W, Costanza ME, Evans III WP, Foster Jr RS, Hendrick E, Eyre HJ, Sener S: American Cancer Society guidelines for breast cancer screening: update 2003. CA: a cancer journal for clinicians 2003, 53(3):141–169.

7. Ali AN, Yuan FJ, Ying CH, Ahmed NZ: Effectiveness of intervention on awareness and knowledge of breast self-examination among the potentially at risk population for breast cancer. Asian Oncology Research Journal 2019:1–13.

8. Gözüm S, Karayurt Ö, Kav S, Platin N: Effectiveness of peer education for breast cancer screening and health beliefs in eastern Turkey. Cancer nursing 2010, 33(3):213–220.

9. Bala D, Gameti H: An educational intervention study of breast self examination (BSE) in 250 women beneficiaries of urban health centers of west Zone of Ahmedabad. Healthline 2011, 2(2):46–49.

10. Ginsburg OM, Chowdhury M, Wu W, Chowdhury MTI, Pal BC, Hasan R, Khan ZH, Dutta D, Saeem AA, Al-Mansur R: An mHealth model to increase clinic attendance for breast symptoms in rural Bangladesh: can bridging the digital divide help close the cancer divide? The oncologist 2014, 19(2):177.

11. Islam RM, Bell RJ, Billah B, Hossain MB, Davis SR: Awareness of breast cancer and barriers to breast screening uptake in Bangladesh: A population based survey. Maturitas 2016, 84:68–74.

12. Amin MN, Uddin MG, Uddin MN, Rahaman MZ, Siddiqui SA, Hossain MS, Islam MR, Hasan MN, Uddin SN: A hospital based survey to evaluate knowledge, awareness and perceived barriers regarding breast cancer screening among females in Bangladesh. Heliyon 2020, 6(4):e03753.

13. Ahmed MS, Sayeed A, Mallick T, Syfuddin H: Knowledge and Practices on Breast Cancer among Bangladeshi Female University Students: A Cross-sectional Study. Asian Pacific Journal of Cancer Care 2020, 5(1):19–25.

14. Tithi NS, Asaduzzaman M, Nasrin N, Monjur M: A Cross-sectional Survey on Knowledge regarding Breast Cancer and Breast Self-examination among Bangladeshi Women. Breast cancer 2018, 236(26.17):22.45.

15. Oeffinger KC, Fontham ET, Etzioni R, Herzig A, Michaelson JS, Shih Y-CT, Walter LC, Church TR, Flowers CR, LaMonte SJ: Breast cancer screening for women at average risk: 2015 guideline update from the American Cancer Society. Jama 2015, 314(15):1599–1614.

16. Yip CH, Anderson BO: The Breast Health Global Initiative: clinical practice guidelines for management of breast cancer in low-and middle-income countries. Expert review of anticancer therapy 2007, 7(8):1095–1104.

17. Anderson BO: Global Summit Consensus Conference on International Breast Health Care: guidelines for countries with limited resources. The breast journal 2003, 9:S40–41.

18. Huguley Jr CM, Brown RL, Greenberg RS, Clark WS: Breast self-examination and survival from breast cancer. Cancer 1988, 62(7):1389–1396.

19. Abd El Aziz HM, Akl OA, Ibrahim HK: Impact of a health education intervention program about breast cancer among women in a semi-urban area in Alexandria, Egypt. J Egypt Public Health Assoc 2009, 84(1-2):219–243.

20. Rezaeian M, Sharifirad G, Mostafavi F, Moodi M, Abbasi MH: The effects of breast cancer educational intervention on knowledge and health beliefs of women 40 years and older, Isfahan, Iran. Journal of education and health promotion 2014, 3.

21. Ceber E, Turk M, Ciceklioglu M: The effects of an educational program on knowledge of breast cancer, early detection practices and health beliefs of nurses and midwives. Journal of Clinical Nursing 2010, 19(15-16):2363–2371.

22. Yilmaz M, Sayin Y, Cengiz HÖ: The effects of training on knowledge and beliefs about breast cancer and early diagnosis methods among women. European journal of breast health 2017, 13(4):175.

23. Beydağ KD, Yürügen B: The effect of breast self-examination (BSE) education given to midwifery students on their knowledge and attitudes. Asian Pac J Cancer Prev 2010, 11(6):1761–1764.

24. Mazloumi S, Zare M, Feisal M, Maleki F, Servat F, Ahmadieh M: Effects of health education on knowledge, attitude and practice of female teachers in Yazd intermediate schools on breast cancer. 2006.

25. Öztürk M, Engin V, Kişioğlu A, Yilmazer G: Effects of education on knowledge and attitude of breast self examination among 25+ years old women. Eastern Journal of Medicine 2000, 5(1):13–17.

