## Additional file for "Effectiveness of educational intervention on breast cancer knowledge and breast self-examination among female university students in Bangladesh: a pre-post quasi-experimental one group study"

**Table S1:** Knowledge about symptoms of breast cancer

| Variables | Pre-test | | Post-test | | Percent of correct changes | Mc-Nemar test *p*-vale |
|---|---|---|---|---|---|---|
| | n | % | n | % | | |
| **Sagging of breast (neg question)** | | | | | | |
| No | 121 | 30.3 | 304 | 76.0 | 45.7 | <0.001 |
| Yes | 102 | 25.5 | 41 | 10.3 | | |
| Don't know | 177 | 44.3 | 55 | 13.8 | | |
| **Nipple discharge other than breast milk including blood or pus** | | | | | | |
| Yes | 119 | 29.8 | 310 | 77.5 | 47.7 | <0.001 |
| No | 73 | 18.3 | 32 | 8.0 | | |
| Don't know | 208 | 52.0 | 58 | 14.5 | | |
| **Swelling of part of the breast** | | | | | | |
| Yes | 136 | 34.0 | 292 | 73.0 | 39 | <0.001 |
| No | 69 | 17.3 | 35 | 8.8 | | |
| Don't know | 195 | 48.8 | 73 | 18.3 | | |
| **Shrinking of breast skin** | | | | | | |
| Yes | 132 | 33.0 | 319 | 79.8 | 46.8 | <0.001 |
| No | 84 | 21.0 | 28 | 7.0 | | |
| Don't know | 184 | 46.0 | 53 | 13.3 | | |
| **Changes in breast shape and size** | | | | | | |
| Yes | 161 | 40.3 | 323 | 80.8 | 40.5 | <0.001 |
| No | 47 | 11.8 | 22 | 5.5 | | |
| Don't know | 192 | 48.0 | 55 | 13.8 | | |
| **Color change of breast including redness or flaky skin** | | | | | | |
| Yes | 193 | 48.3 | 335 | 83.8 | 35.5 | <0.001 |
| No | 33 | 8.3 | 22 | 5.5 | | |
| Don't know | 174 | 43.5 | 43 | 10.8 | | |
| **New lump in the breast or armpit** | | | | | | |
| Yes | 171 | 42.8 | 328 | 82.0 | 39.2 | <0.001 |
| No | 25 | 6.3 | 25 | 6.3 | | |
| Don't know | 204 | 51.0 | 47 | 11.8 | | |
| **Abnormal pain in breast** | | | | | | |
| Yes | 162 | 40.5 | 329 | 82.3 | 41.8 | <0.001 |
| No | 49 | 12.3 | 34 | 8.5 | | |
| Don't know | 189 | 47.3 | 37 | 9.3 | | |

**Table S2:** Knowledge about risks of breast cancer

| Variables | Pre-test | | Post-test | | Percent of correct changes | Mc-Nemar test *p*-vale |
|---|---|---|---|---|---|---|
| | n | % | n | % | | |
| **Not feeding breast milk to children** | | | | | | |
| Yes | 83 | 20.8 | 309 | 77.3 | 56.5 | <0.001 |
| No | 92 | 23.0 | 39 | 9.8 | | |
| Don't know | 225 | 56.3 | 52 | 13.0 | | |
| **Not being physically active** | | | | | | |
| Yes | 173 | 43.3 | 307 | 76.8 | 33.5 | <0.001 |
| No | 84 | 21.0 | 38 | 9.5 | | |
| Don't know | 143 | 35.8 | 55 | 13.8 | | |
| **Previous treatment with hormones or radiation** | | | | | | |
| Yes | 117 | 29.3 | 306 | 76.5 | 47.2 | <0.001 |
| No | 63 | 15.8 | 29 | 7.2 | | |
| Don't know | 220 | 55.0 | 65 | 16.3 | | |
| **Food habit** | | | | | | |
| Yes | 143 | 35.8 | 295 | 73.8 | 38 | <0.001 |
| No | 57 | 14.2 | 19 | 4.8 | | |
| Don't know | 200 | 50.0 | 86 | 21.5 | | |
| **Cyst in breast** | | | | | | |
| Yes | 122 | 30.5 | 309 | 77.3 | 46.8 | <0.001 |
| No | 69 | 17.3 | 29 | 7.2 | | |
| Don't know | 209 | 52.3 | 62 | 15.5 | | |
| **Genetic reasons/family history** | | | | | | |
| Yes | 117 | 29.3 | 313 | 78.3 | 49 | <0.001 |
| No | 68 | 17.0 | 17 | 4.3 | | |
| Don't know | 215 | 53.8 | 70 | 17.5 | | |
| **Alcohol consumption** | | | | | | |
| Yes | 145 | 36.3 | 296 | 74.0 | 37.7 | <0.001 |
| No | 60 | 15.0 | 39 | 9.8 | | |
| Don't know | 195 | 48.8 | 65 | 16.3 | | |
| **Ageing/getting older** | | | | | | |
| Yes | 144 | 36.0 | 262 | 65.5 | 29.5 | <0.001 |
| No | 59 | 14.8 | 37 | 9.3 | | |
| Don't know | 197 | 49.3 | 101 | 25.3 | | |
| **Consuming birth control pill regularly** | | | | | | |
| Yes | 147 | 36.8 | 294 | 73.5 | 36.7 | <0.001 |
| No | 44 | 11.0 | 37 | 9.3 | | |
| Don't know | 209 | 52.3 | 69 | 17.3 | | |
| **Obesity** | | | | | | |
| Yes | 149 | 37.3 | 334 | 83.5 | 46.2 | <0.001 |
| No | 10 | 2.5 | 9 | 2.3 | | |
| Don't know | 241 | 60.3 | 57 | 14.2 | | |

**Table S3:** Knowledge about treatment of breast cancer

| Variables | Pre-test n | Pre-test % | Post-test n | Post-test % | Percent of correct changes | Mc-Nemar test *p*-vale |
|---|---|---|---|---|---|---|
| **Breast cancer is curable if detected at early stage** | | | | | | |
| Yes | 144 | 36.0 | 326 | 81.5 | 45.5 | <0.001 |
| No | 48 | 12.0 | 14 | 3.5 | | |
| Don't know | 208 | 52.0 | 60 | 15.0 | | |
| **Chemotherapy is an effective treatment of breast cancer** | | | | | | |
| Yes | 161 | 40.3 | 283 | 70.8 | 30.5 | <0.001 |
| No | 58 | 14.5 | 30 | 7.5 | | |
| Don't know | 181 | 45.3 | 87 | 21.8 | | |
| **Surgery is an effective treatment of breast cancer** | | | | | | |
| Yes | 149 | 37.3 | 318 | 79.5 | 42.2 | <0.001 |
| No | 53 | 13.3 | 31 | 7.8 | | |
| Don't know | 198 | 49.5 | 51 | 12.8 | | |
| **Hormonal therapy is an effective treatment of breast cancer** | | | | | | |
| Yes | 111 | 27.8 | 304 | 76.0 | 48.2 | <0.001 |
| No | 37 | 9.3 | 37 | 9.3 | | |
| Don't know | 252 | 63.0 | 59 | 14.8 | | |
| **Curable by Alternative medicines (neg question)** | | | | | | |
| No | 68 | 17.0 | 316 | 79.0 | 62 | <0.001 |
| Yes | 69 | 17.3 | 51 | 12.8 | | |
| Don't know | 263 | 65.8 | 33 | 8.3 | | |
| **Curable by herbal treatment (neg question)** | | | | | | |
| No | 82 | 20.5 | 304 | 76.0 | 55.5 | <0.001 |
| Yes | 86 | 21.5 | 46 | 11.5 | | |
| Don't know | 232 | 58.0 | 50 | 12.5 | | |

**Table S4:** Knowledge about prevention of breast cancer

| Variables | Pre-test | | Post-test | | Percent of correct changes | Mc-Nemar test *p*-vale |
|---|---|---|---|---|---|---|
| | n | % | n | % | | |
| **Breast cancer is 100% preventable (neg question)** | | | | | | |
| No | 128 | 32.0 | 314 | 78.5 | 46.5 | <0.001 |
| Yes | 99 | 24.8 | 52 | 13.0 | | |
| Don't know | 173 | 43.3 | 34 | 8.5 | | |
| **Feeding breastmilk to child regularly** | | | | | | |
| Yes | 134 | 33.5 | 292 | 73.0 | 39.5 | <0.001 |
| No | 58 | 14.5 | 16 | 4.0 | | |
| Don't know | 208 | 52.0 | 92 | 23.0 | | |
| **Not wearing underwear all the time** | | | | | | |
| Yes | 93 | 23.3 | 324 | 81.0 | 57.7 | <0.001 |
| No | 47 | 11.8 | 23 | 5.8 | | |
| Don't know | 260 | 65.0 | 53 | 13.3 | | |
| **Early detection by BSE and clinical examination** | | | | | | |
| Yes | 84 | 21.0 | 323 | 80.8 | 59.8 | <0.001 |
| No | 23 | 5.8 | 5 | 1.3 | | |
| Don't know | 293 | 73.3 | 72 | 18.0 | | |
| **Early detection and seeking medical assistance if any symptoms are found** | | | | | | |
| Yes | 288 | 72.0 | 331 | 82.8 | 10.8 | 0.001 |
| No | 19 | 4.8 | 9 | 2.3 | | |
| Don't know | 93 | 23.3 | 60 | 15.0 | | |
| **Maintaining ideal body weight** | | | | | | |
| Yes | 156 | 39.0 | 334 | 83.5 | 44.5 | <0.001 |
| No | 68 | 17.0 | 19 | 4.8 | | |
| Don't know | 176 | 44.0 | 47 | 11.8 | | |
| **Being physically active** | | | | | | |
| Yes | 241 | 60.3 | 314 | 78.5 | 18.2 | <0.001 |
| No | 30 | 7.5 | 23 | 5.8 | | |
| Don't know | 129 | 32.3 | 63 | 15.8 | | |
| **Vaccine (neg question)** | | | | | | |
| No | 200 | 50.0 | 297 | 74.3 | 24.3 | <0.001 |
| Yes | 5 | 1.3 | 14 | 3.5 | | |
| Don't know | 195 | 48.8 | 89 | 22.3 | | |
| **Food habit** | | | | | | |
| Yes | 204 | 51.0 | 327 | 81.8 | 30.8 | <0.001 |
| No | 47 | 11.8 | 20 | 5.0 | | |
| Don't know | 149 | 37.3 | 53 | 13.3 | | |

**Table S5:** Knowledge about screening of breast cancer

| Variables | Pres-test | | Post-test | | Percent of correct changes | Mc-Nemar test *p*-vale |
|---|---|---|---|---|---|---|
| | n | % | n | % | | |
| **Clinical examination is a type of screening** | | | | | | |
| Yes | 109 | 27.3 | 294 | 73.5 | 46.2 | <0.001 |
| No | 125 | 31.3 | 22 | 5.5 | | |
| Don't know | 166 | 41.5 | 84 | 21.0 | | |
| **Mammography is a type of screening** | | | | | | |
| Yes | 160 | 40.0 | 282 | 70.5 | 30.5 | <0.001 |
| No | 65 | 16.3 | 25 | 6.3 | | |
| Don't know | 175 | 43.8 | 93 | 23.3 | | |
| **Breast self-examination is a type of screening** | | | | | | |
| Yes | 168 | 42.0 | 349 | 87.3 | 45.3 | <0.001 |
| No | 42 | 10.5 | 6 | 1.5 | | |
| Don't know | 190 | 47.5 | 45 | 11.3 | | |
| **Ultrasound is a type of screening** | | | | | | |
| Yes | 134 | 33.5 | 341 | 85.3 | 51.8 | <0.001 |
| No | 67 | 16.8 | 10 | 2.5 | | |
| Don't know | 199 | 49.8 | 49 | 12.3 | | |
| **Biopsy is a type of screening** | | | | | | |
| Yes | 156 | 39.0 | 326 | 81.5 | 42.5 | <0.001 |
| No | 46 | 11.5 | 10 | 2.5 | | |
| Don't know | 198 | 49.5 | 64 | 16.0 | | |

**Table S6:** Knowledge about process of breast cancer

| **Variables** | Pre-test | | Post-test | | Percent of correct changes | Mc-Nemar test *p*-vale |
|---|---|---|---|---|---|---|
| | n | % | n | % | | |
| **Inspecting breast visually Infront of a mirror to look for any changes like size, shape, color, unusual discharge or nipple inversion** | | | | | | |
| Yes | 127 | 31.8 | 312 | 78.0 | 46.2 | <0.001 |
| No | 139 | 34.8 | 51 | 12.8 | | |
| Don't know | 134 | 33.5 | 37 | 9.3 | | |
| **Inspecting breast and look for changes Infront of mirror by holding arms at sides, by arms over head, by hands on hips and tighten chest muscle, and by bending forward with hands on hips** | | | | | | |
| Yes | 128 | 32.0 | 328 | 82.0 | 50 | <0.001 |
| No | 144 | 36.0 | 44 | 11.0 | | |
| Don't know | 128 | 32.0 | 28 | 7.0 | | |
| **Inspecting breast by lying down on back with pillow under shoulder and use pads of three middle fingers to give pressure in circle, up and down pattern for each breast** | | | | | | |
| Yes | 126 | 31.5 | 320 | 80.0 | 48.5 | <0.001 |
| No | 119 | 29.8 | 34 | 8.5 | | |
| Don't know | 155 | 38.8 | 46 | 11.5 | | |
| **Feel for changes in armpits by fingers in up down vertical** | | | | | | |
| Yes | 123 | 30.8 | 324 | 81.0 | 50.2 | <0.001 |
| No | 118 | 29.5 | 41 | 10.3 | | |
| Don't know | 159 | 39.8 | 35 | 8.8 | | |
| **Inspecting breasts while bathing with soap** | | | | | | |
| Yes | 125 | 31.3 | 292 | 73.0 | 41.7 | <0.001 |
| No | 108 | 27.0 | 46 | 11.5 | | |
| Don't know | 167 | 41.8 | 62 | 15.5 | | |
